# Anatomy-informed multimodal learning for myocardial infarction prediction

**DOI:** 10.1101/2023.07.11.23292509

**Authors:** Ivan-Daniel Sievering, Ortal Senouf, Thabo Mahendiran, David Nanchen, Stephane Fournier, Olivier Muller, Pascal Frossard, Emmanuel Abbé, Dorina Thanou

**Affiliations:** Swiss Data Science Center, ETH Zurich and EPFL, Switzerland; Signal Processing Laboratory 4, and the Chair of Mathematical Data Science EPFL, Switzerland; Chair of Mathematical Data Science EPFL; Unisante, Switzerland; Department of Cardiology, CHUV, Switzerland; Signal Processing Laboratory 4, EPFL, Switzerland; Chair of Mathematical Data Science EPFL, Switzerland; Center for Intelligent Systems, EPFL, Switzerland

**Keywords:** Coronary Artery Disease, Myocardial Infarction, Invasive Coronary Angiography, Diameter Stenosis, Deep Learning, Multimodal data, AI for cardiology

## Abstract

In patients with coronary artery disease the prediction of future cardiac events such as myocardial infarction (MI) remains a major challenge. In this work, we propose a novel anatomy-informed multimodal deep learning framework to predict future MI from clinical data and Invasive Coronary Angiography (ICA) images. The images are analyzed by Convolutional Neural Networks (CNNs) guided by anatomical information, and the clinical data by an Artificial Neural Network (ANN). Embeddings from both sources are then merged to provide a patient-level prediction. The performance of our framework on a clinical study of 445 patients admitted with acute coronary syndromes confirms that multimodal learning increases the predictive power and achieves good performance, which outperforms the prediction obtained by each modality independently as well as that of interventional cardiologists. To the best of our knowledge, this is the first attempt towards combining multimodal data through a deep learning framework for future MI prediction.

## 1 Introduction

Coronary artery disease (CAD), a leading cause of death worldwide [1], refers to a disease of the coronary arteries that supply blood to the heart muscle. The disease results, primarily, from the development of plaques of atherosclerosis in the arterial wall, which ultimately lead to narrowings (stenoses) and reduced blood flow. In the acute setting, CAD takes the form of an acute coronary syndrome (ACS), with the most feared manifestation being a myocardial infarction (MI) resulting from the rupture of a plaque of atherosclerosis and the subsequent, abrupt interruption of coronary blood flow. The resulting necrosis of the heart muscle can lead to numerous complications including a reduction in heart function (heart failure), arrhythmia (including cardiac arrest), and death.

However, CAD is a complex pathological process with numerous factors that drive its development, its progression, and its risk of provoking an MI. At a local level, the diameter of the stenosis does correlate with the risk of MI, but it remains insufficient as a predictor of MI, as highlighted by the influence of other local factors such the hemodynamic impact of the stenosis [2]. At a patient level, important drivers of CAD include cardiovascular risk factors (e.g., age, sex, hypertension, diabetes, dyslipidemia), all of which can influence the risk of MI. As a result, in clinical practice, future MI prediction remains a challenge. This is highlighted by the fact that up to 10% of patients with stenoses deemed non-significant (i.e., stenoses *<* 50% without a significant hemodynamic impact) still present an MI or a need for urgent revascularization (i.e., invasive treatment such as stenting) in the ensuing two years [3]. Even among patients treated for an MI, the risk of short- and long-term adverse outcomes remains significant. Reinfarction represents a significant cause of poor outcomes among MI patients, with rates as high as 4% at one year and 7% at three years [4].

In both acute and chronic settings, invasive coronary angiography (ICA) remains the gold standard investigation for the diagnosis and evaluation of CAD in clinics. ICA involves continuous X-ray (i.e., fluoroscopy) with simultaneous injection of radiopaque contrast into the coronary arteries, thus permitting the identification of coronary stenoses. In current clinical practice, stenosis severity is still often only determined by the physician’s estimation of percentage reduction in arterial diameter. With this approach, a diameter stenosis *≥* 70% is generally considered a strong indicator of a clinically significant lesion, and thus a criterion for treatment (e.g., coronary stenting). New innovative approaches are thus needed to drive progress in this field.

Recent advances in machine learning (ML), in combination with the availability of multimodal data, show promise for capturing and quantifying the complexity of CAD. Some works have already focused on predicting MI in the next months from patients’ clinical data using traditional ML algorithms such as Logistic Regression, Random Forest, Gradient Boosting, (e.g., [5], [6]), with limited success. At the same time, convolutional neural networks (CNNs) have been successfully applied in detecting stenoses and inferring their severity directly from ICA images [7], but without tackling the challenge of future event prediction. First step towards future MI prediction from ICA images was performed in [8] and [9], where a deep learning framework was able to provide significant gains in predicting future culprit lesions, i.e., lesions that lead to future events. This study however was performed at a lesion level and on patients with stable coronary disease. Given the complexity of CAD and the numerous influencers of the risk of future MI, a multimodal approach that extracts knowledge from all available patient data appears crucial for predicting future events.

In this work, we depart from the task of lesion-level MI prediction from ICA images, instead tackling the challenge of patient-level MI prediction using both ICA and clinical information. In particular, we propose an anatomy-informed deep learning framework that combines ICA imaging views from the three different arteries of the coronary tree (see Fig. 1 for an example of the different arteries), cardiologist guidance on significant anatomical regions, and clinical data in order to predict the occurrence of future MI in patients presenting with an acute coronary syndrome at baseline. The problem is particularly challenging for ML settings not only due to the systemic and biological complexity of the disease, but also due to the frequency of the disease. Due to its invasive nature, patients undergo ICA only if there is significant justification. Moreover, out of all these patients, only a small percentage will experience an MI in the future, limiting further the number of MI events. This low and unbalanced data regime is particularly challenging for ML algorithms.

**Fig. 1.**
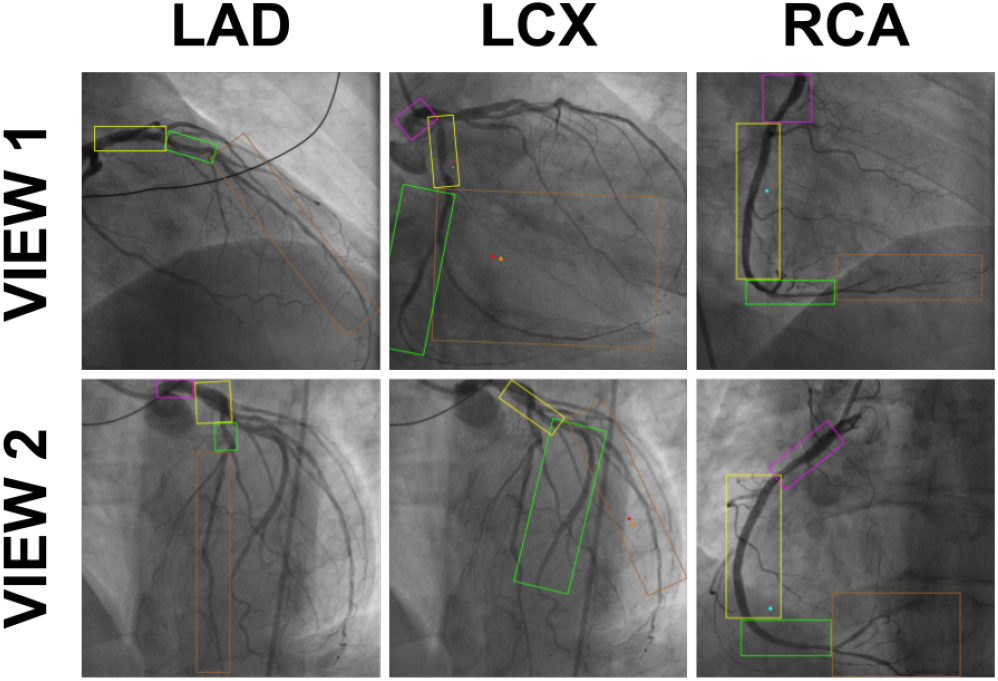
Annotated ICA images of a patient; two views for each of three different arteries. The boxes indicate different anatomical segments and the red dots that the segment is responsible of future MI.

To partially overcome these issues, we exploit our knowledge of the coronary artery tree, and cast the problem as a multi-objective learning framework, with the goal to predict MI both at an artery (auxiliary task) and at a patient level (main task). The MI prediction at an artery level is achieved by learning discriminative features from the corresponding artery views using anatomy-informed CNNs, combined with learned representations of the clinical data, computed by an ANN. These jointly learned artery-specific representations are then concatenated to obtain a prediction at a patient level, which is provided by another ANN. The results obtained in a clinical cohort of 445 patients confirm that considering jointly both image modalities and clinical data (AUC: 0.67 *±* 0.04 & F1-Score: 0.36 ± 0.12) provides significantly better predictive power than learning from each modality independently (only using ICA images: AUC: 0.64 ± 0.04 & F1-Score: 0.30 ± 0.11 and only using clinical data: AUC: 0.63 ± 0.04 & F1-Score: 0.28 ± 0.06). The multimodal learning scheme also outperforms the prediction of interventional cardiologists from the ICA images (AUC: 0.54 ± 0.04 & F1-Score: 0.18 ± 0.04). To the best of our knowledge, this is the first attempt towards providing a global prediction score for future coronary artery events by exploiting, in a data-driven manner, conjointly the patients’ clinical data and their ICA images.

The rest of the paper is organized as follows. First, we provide a description of the clinical study, the different data modalities, and the cardiologist-guided annotation of the anatomical segments of the coronary artery tree. Then, we present our novel multi-objective learning approach for predicting MI from multimodal data. Finally, we illustrate the performance of our framework on the clinical study, and compare it with different baselines that rely on learning from a single modality or inferring from interventional cardiologist expertise.

## II Clinical study

The SPUM-ACS (Special Program University Medicine - Acute Coronary Syndromes) registry is a cohort of consecutive patients admitted with acute coronary syndromes (MI or unstable angina) to four university hospitals in Switzerland between 2009 and 2017. Further details of this registry have been reported previously [10]. For the present study, patients hospitalised with ICA images available for analysis are included. The clinical endpoint considered in this work is MI, i.e., we consider patients who had an MI in the next five years after the acute event (the acute coronary syndrome at baseline). Our approach could be generalized to other datasets that include enough ICA views and similar clinical information about the patient.

### A. ICA images

The dataset consists of clinical data and ICA images of 445 patients, out of which 47 experienced an MI during the followup period. Six ICA images were extracted from the baseline ICA of each patient: three arteries (left anterior descending (LAD), left circumflex (LCX) and right coronary artery (RCA)) viewed from two angles with *≈*30° of difference. An illustrative example is shown in Fig. 1. Most of the ICA images are 1524×1524 pixels in size. Those that have different sizes are cropped or interpolated to this size. Only patients with at least one view of each artery are considered. In order to have a fixed number of views per patient, for patients with only one view, we consider that view twice, while for patients with more than two views, we randomly select two views per patient. The initial number of patients in the dataset is 709, but for 264 of them, the dataset does not contain at least one view of each artery, or contain invalid values, and thus only 445 patients are considered.

### B. Cardiology-guided annotation of anatomical segments

Each of the views is annotated by an interventional cardiologist into anatomical segments as defined by the SYNTAX system [11] (colored boxes in Fig. 1, with the color code being consistent across patients). Each segment is then labeled as being responsible for future MI or not (red dots in Fig. 1). The anatomical segments will be used as attention masks in order to guide the learning of the proposed algorithm towards important regions of the coronary artery tree.

### C. Clinical data

The clinical data consists of sex, age, body mass index, diabetes, smoking, hypertension, hypercholesterolaemia, previous cardiovascular disease, Killip class, previous cardiac arrest and kidney function, which are known to be cardiovascular risk factors. The non-categorical data are normalised (mean subtracted and divided by the standard deviation). Each missing data is replaced by the median, if it belongs to a continuous column (i.e., values of the column can take any value in a given range), or by the most frequent value, if it belongs to a categorical column (i.e., values of the column come from a given set of possible values).

## III A multimodal learning framework for mi prediction

### A. Proposed model

We propose a novel, anatomy-informed learning framework that predicts MI by combining coronary artery anatomical information from ICA images with patient-level clinical data, e.g., cardiovascular risk factors. Our approach takes the images of the different ICA views of the artery as an input along with the patient’s clinical data, permitting the prediction of MI at an artery level. These artery-level predictions are then combined in order to predict whether the patient will have an MI. A summary of the architecture is illustrated in Fig. 3. In what follows, we elaborate on each building block of the architecture, and each data modality.

### Imaging data

To extract predictive features from ICA images, we adapt state-of-the-art image-based deep learning architectures to the specificity of the coronary artery images. For each main artery of the coronary tree, we propose a CNN-based architecture, which receives as input the two views of the artery as well as the corresponding attention masks that indicate the bounding boxes drawn by the physicians. These masks help the network focus on the main anatomical segments of the coronary artery tree and not on the background, see Fig. 2. Given that both views represent a snapshot of the same 3D artery from different angles in the 2D space, we use this anatomy-informed fact to process them jointly through two siamese networks as we expect that similar features are present in both pictures. The backbone is a ResNet-18 network [12], which is a well-known state-of-the-art model. We then achieve a global representation of the ICA images by concatenating the representations of both views, followed by average pooling, flattening and dropout, in order to reduce the dimension of the embedded space and to diminish overfitting. Finally, this global representation of the views is given as an input to a classification layer, which is responsible for predicting MI in each artery only from imaging data. This global representation will be used for the multimodal prediction, jointly with the embedding of the clinical data.

**Fig. 2.**
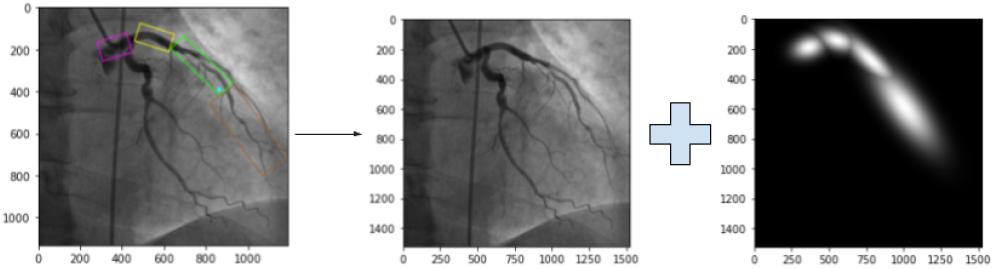
The annotated ICA images (left) are converted to a raw image (center) and a mask (right) that indicates the different anatomical segments. The mask is created by generating Gaussian functions centered on the sections’ rectangle and using the same width and height.

**Fig. 3.**
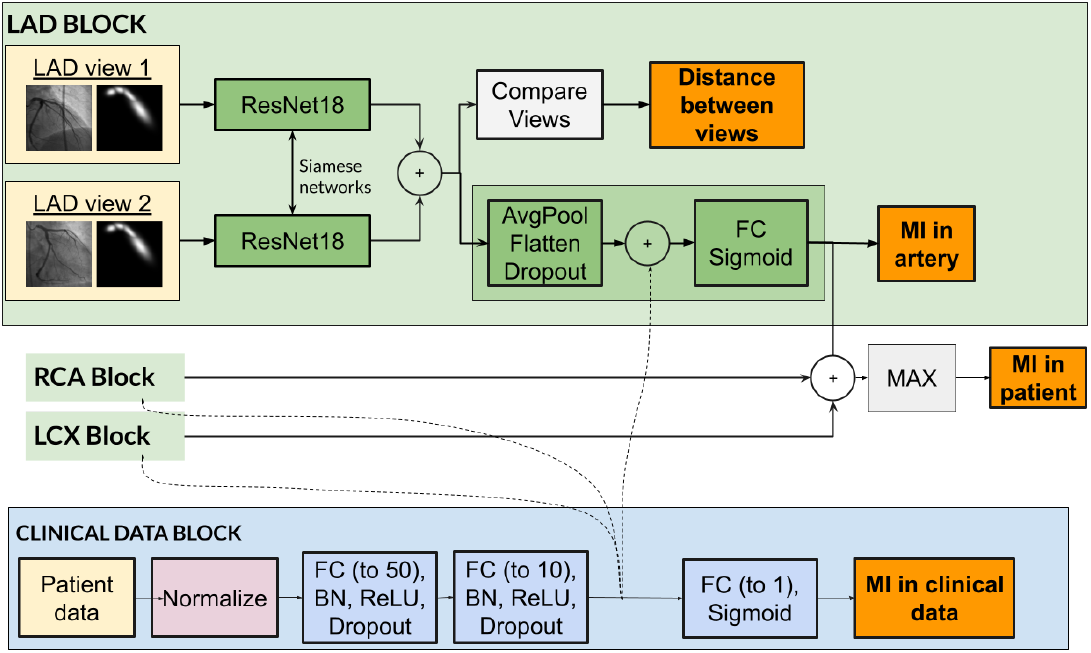
Anatomy-informed multimodal framework for MI prediction. The patient data is processed by the clinical data block and the two views of each artery are processes by artery blocks. A prediction is provided for each artery and the prediction at patient level is the maximum of these three predictions. Note that the decision at the artery level is influenced by the clinical data.

### Clinical data

In parallel, clinical data are processed through two hidden layers of 50 and 10 neurons (Fully connected (FC) layer, batch normalisation, ReLU activation, dropout), inspired by [13]. The feature representation extracted from this pipeline defines the embedding of the clinical data. This embedding is given as an input to a classification layer (FC, Sigmoid), which is responsible for predicting patient level MI only from clinical data, which provides another auxiliary loss. This embedding will also be used for the multimodal prediction, jointly with the images’ embedding.

### Patient-level prediction (multimodal model)

Once ICA and clinical data have been processed independently, their representations are combined in order to achieve a multimodal embedding per artery. We follow a strategy similar to [14] and [15], by concatenating the representations of the ICA images and the clinical data, and further analysing them through a FC layer activated by a Sigmoid function. This output is used to compute the probability of an MI for a given artery. Finally, the prediction at the patient level is defined as the worst-case prediction of the three arteries, i.e., a single MI prediction at an artery level is enough to predict MI at a patient level. The independent prediction for each artery is motivated by the anatomy-informed fact that MI is often a local event, i.e., information from an artery does not necessarily improve the prediction in another one.

### B. Training procedure

Our multimodal network is trained by minimizing a loss function (*l*) that takes into account each of the building blocks mentioned above. In particular, we aim to improve the overall MI predictions by optimizing the artery level predictions from multimodal embeddings (*l*_*pred*(*LAD/LCX/RCA*)_), the patient level predictions from clinical data (*l*_*clinical*_), and the overall patient level prediction from both clinical and artery level embeddings (*l*_*patient*_). In addition, for each artery view, we define an auxiliary loss function that enforces similarity between views in the embedding space (*l*_*simi*(*LAD/LCX/RCA*)_). The loss will be zero if the views share the same information (this loss helps the model to learn the features to detect as we expect that both views mostly contain similar information). This function is set to be the mean Euclidean distance between the features extracted from the two pairs of views of the same artery.

Thus, the complete loss that consists of the above mentioned terms, can be defined as presented in Eq. 1. The weight of the main loss (patient level prediction) is always fixed to one as it is our main target and the others (*w* = [*w*_1_, *w*_2_, *w*_3_]) are considered as hyperparameters (ranging from 0 to 1).

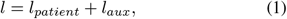

where

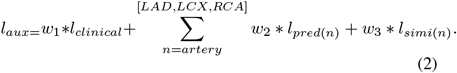

Due to the imbalanced nature of our dataset (47 MI out of 445), the classification loss is defined as an AUC-specific loss function ([16], optimized by PESG [17]). To tackle the imbalance, the MI samples of the dataset are oversampled through data augmentation. Different image-based augmentation methods are applied to ICA images in the training set (no data augmentation is applied to the clinical data). Each data augmentation technique has a given probability of happening, and is applied sequentially to the images in the following order:

1. Random cropping (probability of happening: 20%): a subimage is cropped on the image, which can have a ratio between 75% and 125% of the original image and between 80% and 99% of the size of the image. The image is then resized to the input size;
2. Random rotation (probability of happening: 20%): the image is rotated between -30° and +30°;
3. Color (probability of happening: 20%): brightness is altered between 80% and 120%, contrast between 80% and 120%, saturation between 80% and 120% and hue between -20% and 20%.

For the cropping and rotation, the same values are applied to the raw image and its mask (otherwise, the mask does not provide the correct insight). The parameters’ values have been fixed heuristically.

The weights of the ResNet-18 backbones are initialized with a pre-trained network on ImageNet [18] (it was found that pre-trained models reach better performance, despite the fact that our images are not “natural” images). The models are trained using 5-fold cross-validation on the training set, with similar number of MI patients in each fold. The multimodal network is trained during 20 epochs with PESG (gamma: 595, margin: 0.99) and a batch size of 4. The weight decay and the dropout are fixed to 0.003, and 31% respectively. The learning rate is initially set to 0.077, and a scheduler divides by 10 the learning at each plateau (3 consecutive epochs without metric improvement). The weight of artery loss, distance loss, and clinical data loss are set to 0.06, 0.007 and 0.006 respectively. All these hyperparameters have been selected using grid search.

## IV Results

In this section, we evaluate our multimodal frameworks for MI prediction on the clinical study presented in Section II. First, we introduce our baseline methods that are based on predicting from each modality independently, as well as a comparison with the prediction score achieved from the visual inspections of interventional cardiologists. Next, the obtained results are discussed.

### A. Baselines

#### a) Single modality: ICA

We truncate the multimodal model so that it uses only the ICA images, i.e., without the “Clinical Data Block” in Fig. 3. We compute predictions per artery and define the patient level prediction as the worst-case scenario from each artery. The model is trained using the AUC-loss (as for the multimodal).

The model is trained for 20 epochs with PESG optimizer (gamma: 411, margin: 0.81) and a batch size of 4. The weight decay is set to 0.007 and the dropout to 0.6%. The starting learning rate is 0.03, divided by 10 after 3 epochs without improvement. The weight of the artery loss is 0.002, and the weight of the distance loss 0.0008.

#### b) Single modality: Clinical data

We use only the “Clinical Data Block” from the multimodal model presented in Fig. 3 and thus predict MI only based on clinical data.

The model is trained during 300 epochs with PESG (gamma: 470, margin: 0.92) and a batch size of 32. A Kaiming Normal [19] is used for initialisation. The weight decay is 0.0048, and the dropout is 49.76%. The starting learning rate is 0.004, divided by 10 after 25 epochs without improvement.

#### C) Visual inspection from interventional cardiologists

The exact same set of patients is evaluated by two blinded interventional cardiologists, who analysed the ICA views of each patients and provided their patient-level prediction of MI.

#### d) Naive predictor

A naive strategy is applied: it classifies the sample as positive 10.6% of the time, and otherwise as negative. This probability corresponds to distribution of MI in the dataset (47 MI among 445 patients (10.6%)). Thus, this benchmark shows the performance of random guesses.

## B. Evaluation metrics

The dataset is split in training set and testing set. The performances is computed i) by applying a 5-fold cross validation procedure on the training set and recording the mean and the standard deviation performances of the models on the validation sets, and ii) by training the model on the whole training set and testing it on the testing set.

Four evaluation metrics [20] are considered:

- AUC-ROC measures the balance between the True Positive Rate and the False Positive Rate;
- Precision measures the percentage of True Positive among the samples classified as positive;
- Recall measures the percentage of positive correctly classified;
- F1-Score is the harmonic mean of precision and recall.

## C. Performance Analysis

In Table I, we compare the MI predictive performance of the proposed multimodal framework, as well as the baselines mentioned above during the 5-fold cross validation procedure. We observe that the performance of our multimodal framework is better than learning from each modality independently, highlighting the benefit of extracting predictive features from both.

**TABLE I.** Different modalities: Validation scores (mean±std) obtained from a 5-fold cross-validation on the training set.

| Predictor | F1-Score | AUC-ROC | Precision | Recall |
| --- | --- | --- | --- | --- |
| Naive | 0.10 | 0.50 | 0.10 | 0.10 |
| Interventional Cardiologist 1 | 0.14 | 0.49 | 0.10 | 0.25 |
| Interventional Cardiologist 2 | 0.22 | 0.58 | 0.15 | 0.47 |
| Cardiologists' mean | 0.18 | 0.54 | 0.13 | 0.36 |
| Single modality: Clinical data | 0.28±0.06 | 0.63±0.04 | 0.18±0.04 | **0.72±0.17** |
| Single modality: ICA images | 0.30±0.11 | 0.64±0.04 | 0.21±0.13 | 0.69±0.11 |
| Multimodal framework | **0.36±0.12** | **0.67±0.04** | **0.36±0.18** | 0.44±0.10 |

The performances obtained only from clinical data and only from ICA are similar, highlighting the difficulty of learning directly from ICA. This is most likely attributed to several reasons with the main ones being the limited number of data points, and in particular MI patients, and the important quantity of uninformative background in ICA images. At the same time, the multimodal approach outperforms learning from each of the single modalities independently. These results confirm our intuition that each modality contains different information that, if combined properly, can provide a better predictive score at a patient level. The low predictive performance achieved from expert clinicians indicates the complexity of the task, and the importance of building data-driven tools that could assist clinical decision-making.

Table II shows the performances but by training on the whole training set and testing on the testing dataset. These results have to be considered with great care, the main reason being that the testing dataset contains very few positive cases (5 MI over 89 patients). Moreover, the results are outside of the confidence intervals computed from the validation dataset. However, these results lead to similar conclusions to the ones presented previously.

**TABLE II.** Prediction performance on a test set.

| Predictor | F1-Score | AUC-ROC | Precision | Recall | Specificity |
| --- | --- | --- | --- | --- | --- |
| Interventional Cardiologist 1 | 0.095 | 0.484 | 0.054 | 0.400 | 0.568 |
| Interventional Cardiologist 2 | 0.125 | 0.562 | 0.070 | **0.600** | 0.524 |
| Cardiologists' mean | 0.110 | 0.523 | 0.062 | 0.500 | 0.546 |
| Multimodal framework | **0.167** | **0.627** | **0.100** | **0.600** | **0.654** |

## D. Alternative models

### a) Alternative single modality: ICA

Different variations of the single modality ICA model are considered. First, two losses are used for training, i.e., the AUC-loss (as for the multimodal) and the Binary Cross Entropy (BCE) loss (optimized by Stochastic Gradient Descent (SGD)). Second, two different ways for providing patients predictions are compared: (i) *Max analysis:* We compute predictions per artery and define the patient level prediction as the worst-case scenario from each artery. (ii) *Common:* We concatenate the output of the Siamese networks for all arteries and process the entire feature representation to predict the MI at the patient level. It is represented in Fig. 4. To improve the discriminative power, we add some additional layers in the architecture. Those layers are similar to the ones used in ResNet [12]: max pooling (to reduce the size of the embedding space), a residual convolutional block (two convolutional layers with batch normalization and ReLU activation, connected by a residual connection), average pooling, flattening, dropout, and finally a classification layer activated by Sigmoid. We note that these additional layers increase the total number of parameter of the second model: the *Common* approach has five times more parameters than the *Max* one.

**Fig. 4.**
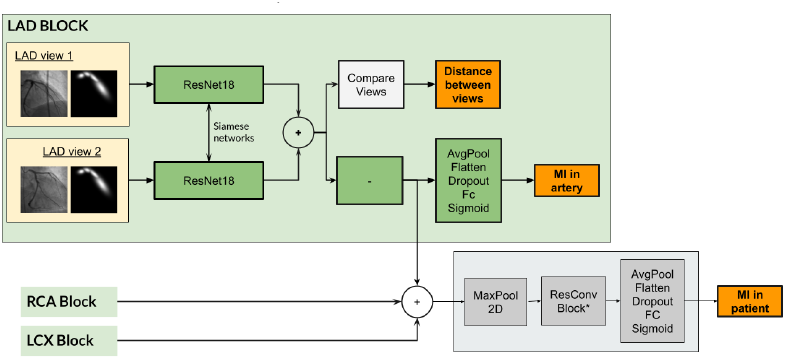
Single modality ICA framework for MI prediction. The two views of each artery are processed separately before being concatenated together into a bigger feature map. This new feature map is further processed through convolutional layers and poolings to finally provide a patient level prediction. The *ResConvBlock* is the set of convolutional blocks connected with skip connections presented in the section IV-A.

In Table III, the different implementations of the single modality models are compared. We notice that the *Max* approaches for the ICA modality (taking the maximum of the prediction of each artery) reaches the same performance as the *Common* one (analyzing all the arteries together) while having much less parameters. Overall, using the AUC loss is better or similar than the BCE loss. For that reason, our solution uses a *Max* architecture trained with AUC-loss. The hyperparameters of those models can be found in Appendix A.

**TABLE III.** Different implementations of single modality: Validation scores (mean±std) obtained from a 5-fold cross-validation on the training set.

| Modality | Implementation | F1-Score | AUC-ROC |
| --- | --- | --- | --- |
| ICA | Max + AUC | 0.30±0.11 | 0.67±0.04 |
| ICA | Max + BCE | 0.28±0.09 | 0.63±0.06 |
| ICA | Common + AUC | 0.30±0.12 | 0.66±0.07 |
| ICA | Common + BCE | 0.31±0.07 | 0.65±0.03 |
| Clinical data | AUC | 0.28±0.06 | - |
| Clinical data | BCE | 0.25±0.05 | - |

### b) Alternative single modality: clinical data

Different traditional ML classifiers have been considered to predict MI from clinical data. Their performance is reported in Table IV. We observe that overall all methods obtain comparable performance. ANN slightly outperforms the others and is more easily implementable in a Neural Network framework but uses significantly more parameters. The ANN is trained with AUC and BCE, and the obtained results are compared in Table III. The hyperparameters of those ANNs can be found in Appendix B. Overall, those results are close to the ones obtained in [5] and [6]. The similarity between the reported performance across different works that predict future outcomes from clinical data, despite using different datasets and models, could be a strong indicator that we may have reached the limits of the predictive capacity of the clinical data, making the use of ICA images on top of clinical data necessary for better predictions.

**TABLE IV.** Different ML algorithm for mi prediction from clinical data: Validation scores (mean±std) obtained from a 5-fold cross-validation on the training set.

| Model | F1-Score<br>(mean±std) | Accuracy<br>(mean±std) (%) |
| --- | --- | --- |
| Decision Tree | 0.23 ± 0.08 | 70.18 ± 0.08 |
| Balanced Random Forest | 0.21 ± 0.11 | 66.23 ± 19.97 |
| Logistic Regression | 0.24 ± 0.10 | 68.37 ± 5.03 |
| Naive Bayes | 0.24 ± 0.09 | 65.63 ± 5.03 |
| Gradient Boosting | 0.24 ± 0.06 | 71.36 ± 2.95 |
| Artificial Neural Network | **0.28 ± 0.06** | **81.12 ± 4.64** |

## V. Discussion

To the best of our knowledge, there are no previous studies that aim to predict a patient’s risk of future MI by combining both image and patient data via a deep learning framework. Thus this pilot study represents the first of its kind and demonstrates the efficacy (albeit modest) of such an approach. From the cardiology perspective, the importance of this specific prediction task cannot be understated. Cardiovascular disease remains the leading cause of the death worldwide [1], despite significant advances in the prevention and treatment of cardiovascular disease in recent years. Even with the application of the best available predictors of cardiovascular risk (e.g. degree of coronary stenosis, hemodynamic impact of a stenosis, risk factors such as diabetes and hypertension), a significant number of patients still go on to experience an MI. This can likely be explained by the complex pathophysiology of coronary artery disease. To tackle this complexity, we propose an approach that integrates pertinent information from different sources.

With regards to the performance of our approach, some average quantitative comparisons can be performed with other works that predict future MI in different settings. Compared to [8], where a deep learning framework was able to predict future culprit lesions from ICA with an F1-score of 0.57, our performance is overall lower. This study was performed at a lesion level and on patients with stable coronary disease. On the one hand, working at a lesion level is a simplified problem for deep learning, as it provides as input the exact lesion, as opposed to the whole artery. At the same time, the clinical cohort of [8] consists of a completely different population, with stable coronary disease, all of which eventually had an MI. In our setup, the cohort consists of patients with acute coronary syndromes, with only 10% suffering from MI in the follow up period. Thus our scenario is more representative of real clinical practice where not all patients will have an MI during follow-up, and the challenge is identifying the ones that will. Compared to algorithms that predict only from clinical data, our results are close to the ones obtained by [5] and [6]. However, while we consider the risk of MI within a follow up period of 5 years using a relatively small dataset (*≈*500 patients) and only 11 clinical features, these works present models developed with significantly larger datasets with many more variables and predict the risk of MI within different time intervals (shorter ones). More specifically, the work in [5] reaches an F1-Score of 0.101 while predicting MI within the next six months for 2 millions patients with 8’000 features. The work of [6] reaches an AUC of 0.72 while predicting MI within 12 months based on 7’000 patients and 192 features. These comparisons suggest that the framework proposed in the current study builds significantly on previous work and has the potential to improve the accuracy of ML-driven predictions of future MI significantly.

Importantly, we recognise that the generalisability of these results to other clinical cohorts needs to be demonstrated due to differing patient populations, as well as variance with respect to the quality and nature of the clinical and imaging data. As such, this study needs to be extended in future work.

## VI Conclusion

In this work, we proposed a multimodal framework based on deep learning, which exploits the knowledge of the main arteries of the coronary tree, and the ICA images corresponding to each of those, as well as clinical patient data in order to predict future MI in patients with acute coronary syndromes. The ICA images are processed by anatomy-guided CNNs, and the clinical patient data is analysed by an ANN. Embeddings from both modalities are then combined to finally provide a patient-level prediction. Experimental results confirmed the superior performance of our method in comparison to learning from each modality separately, but also in comparison to human-based predictions from experienced interventional cardiologists. Although the numerical results of this study should be considered with caution due to the small number of MI patients, the non-trivial performance obtained with the current data is promising. It indicates that the integration, via a well-designed learning framework, of imaging, clinical variables, and clinical expertise (including knowledge of the coronary anatomy), has the potential to improve on current approaches to the highly complex and challenging MI prediction task.

## Data Availability

All data produced in the present work are contained in the manuscript.

## Appendix A Hyperparameters of ICA single modality

In what follows, we document all the hyperparameters of the ICA single modality models. All these parameters are found by grid search.

### A. ICA modality with AUC loss (Max of arteries)

The model was trained for 20 epochs with PESG (gamma: 411, margin: 0.81) and a batch size of 4. The weight decay was set to 0.007 and the dropout to 0.6%. The starting learning rate was 0.03, divided by 10 after 3 epochs without improvement. The weight of the artery loss was 0.002, and the weight of the distance loss 0.0008.

### B. ICA modality with BCE loss (Max of arteries)

Training of the model was done in 20 epochs with SGD (momentum: 0.9) and a batch size of 4. The weight decay was set to 0.015 and the dropout to 0.3%. The starting learning rate was 0.08, divided by 10 after 3 epochs without improvement. The weight of the artery loss was 0.0008, and the weight of the distance loss 0.013.

### C. ICA modality with AUC loss (Common prediction)

The model was trained during 20 epochs with PESG (gamma: 598, margin: 0.90) and a batch size of 4. The weight decay was set to 0.005 and the dropout to 2.9%. The starting learning rate was 0.047, divided by 10 after 3 epochs without improvement. The weight of the artery loss was 0.056, and the weight of the distance loss 0.0066.

### D. ICA modality with BCE loss (Common prediction)

The model was trained during 20 epochs with SGD (momentum: 0.9) and a batch size of 4. The weight decay was set to 0.096 and the dropout to 2.5%. The starting learning rate was 0.01, divided by 10 after 3 epochs without improvement. The weight of the artery loss was 0.057, and the weight of the distance loss 0.096.

## Appendix B Hyperparameters of clinical data single modality

In what follows, we document all the hyperparameters of the clinical data single modality models. All these parameters are found by grid search.

### A. Clinical data modality with AUC loss

The model was trained during 300 epochs with PESG (gamma: 470, margin: 0.92) and a batch size of 32. A Kaiming Normal [19] was used for initialisation. The weight decay was 0.0048, and the dropout was 49.76%. The starting learning rate was 0.004, divided by 10 after 25 epochs without improvement.

### B. Clinical data modality with BCE loss

The model was trained during 300 epochs with SGD (momentum: 0.9) and a batch size of 32. A Kaiming Normal initialised it. The weight decay was 0.0018, and the dropout was 48.36%. The starting learning rate was 0.055, divided by 10 after 25 epochs without improvement.

